# Role of miRNAs induced by oxidized low-density lipoproteins in coronary artery disease: the REGICOR Study

**DOI:** 10.1101/19003749

**Authors:** IR Dégano, I Subirana, N García-Mateo, P Cidad, D Muñoz-Aguado, E Puigdecanet, L Nonell, J Vila, A Camps, F Crepaldi, D de Gonzalo-Calvo, Vc Llorente-Cortés, MT Pérez-García, R Elosua, M Fitó, J Marrugat

**Affiliations:** REGICOR Study Group, IMIM (Hospital del Mar Medical Research Institute), Barcelona, Spain; CIBER of Cardiovascular Diseases (CIBERCV), Instituto de Salud Carlos III (ISCIII), Madrid, Spain; Faculty of Medicine, University of Vic-Central University of Catalonia (UVic-UCC), Vic, Spain; CIBER Epidemiology and Public Health, ISCIII, Madrid, Spain; Department of Biochemistry, molecular biology, and physiology, Institute of Molecular Biology and Genetics (IBGM), University de Valladolid, Spanish National Research Council (CSIC), Valladolid, Spain; Cardiovascular Risk and nutrition Group, IMIM, Barcelona, Spain; CIBER of Obesity and Nutrition (CIBEROBN), ISCIII, Madrid, Spain; MARGenomics, IMIM, Barcelona, Spain; Cardiovascular Research Center, CSIC-ICCC, IBB-Sant Pau, Barcelona, Spain; Cardiovascular epidemiology and genetics Group, IMIM, Barcelona, Spain

**Keywords:** oxidized low-density lipoproteins, miRNAs, acute myocardial infarction, coronary artery disease

## Abstract

**Aims:** Current risk prediction tools are not accurate enough to identify most individuals at high coronary risk. On the other hand, oxidized low-density lipoproteins (ox-LDLs) and miRNAs are actively involved in atherosclerosis. Our aim was to examine the association of ox-LDL-induced miRNAs with coronary artery disease (CAD), and to assess their predictive capacity of future CAD.

**Methods and results:** Human endothelial and vascular smooth muscle cells were treated with oxidized or native LDLs (nLDL), and their miRNA expression was measured with the miRNA 4.0 array, and analyzed with moderated t-tests. Differently expressed miRNAs and others known to be associated with CAD, were examined in serum samples of 500 acute myocardial infarction (AMI) patients and 500 healthy controls, and baseline serum of 117 incident CAD cases and c 485 randomly-selected cohort participants (case-cohort). Both were developed within the REGICOR AMI Registry and population cohorts from Girona. miRNAs expression in serum was measured with custom OpenArray plates, and analyzed with fold change (age and sex-paired case-control) and survival models (case-cohort). Improvement in discrimination and reclassification by miRNAs was assessed. Twenty-one miRNAs were up- or down-regulated with ox-LDL in cell cultures. One of them, 1 (has-miR-122-5p, fold change=4.85) was upregulated in AMI cases. Of the 28 known CAD-associated miRNAs, 11 were upregulated in AMI cases, and 1 (hsa-miR-143-3p, hazard ratio=0.56 [0.38-0.82]) was associated with CAD incidence and improved reclassification.

**Conclusion:** We identified 2 novel miRNAs associated with ox-LDLs (hsa-miR-193b-5p and hsa-miR-1229-5p), and 1 miRNA that improved reclassification of healthy individuals (hsa-miR-143-3p).

## Introduction

Although coronary artery disease (CAD) incidence is decreasing in many European regions [1,2], both the number of first events in low-risk individuals, and the recurrence rate after acute myocardial infarction (AMI) are still high. In general population without cardiovascular disease (CVD), cardiovascular prevention strategies depend on the predicted CVD risk. This risk is calculated with available risk functions, and defines whether the individual receives lifestyle counselling and/or lipid-lowering and hypertension treatments [3]. However, the discrimination of the validated European risk functions ranges between 70-88% [4-8]. This lack of accuracy means that >50% of events occur in individuals classified as with low/moderate risk, which do not qualify for intensive prevention strategies [9]. On the other hand, 18-21% of AMI patients have a second CVD event within 4 years after discharge [10,11]. An improvement in risk prediction in primary and secondary CVD prevention, could decrease the number of events in low risk individuals, and the number of recurrent events in MI patients. But improving risk prediction is not an easy task.

In primary prevention, risk prediction variables have not changed in the last decades. Attempts to improve risk functions have mainly focused on including genetic and/or circulating biomarkers, with little impact on their discrimination and calibration capacity [3]. Discovery of other biomarkers could improve risk stratification, and boost the identification of new biological pathways and therapeutic targets.

MicroRNAs (miRNAs) are potential biomarkers and therapeutic targets. miRNAs allow temporal regulation of gene expression [12] and control essential processes in atherosclerosis and after an AMI [13]. In addition, miRNAs are relatively stable and easy to measure in serum [14].

In the last years, a few studies have identified miRNAs that are associated with better discrimination of high risk individuals in general population [15] and after an AMI with ST segment elevation [16]. However, previous studies examined panels of miRNAs without taking into account key risk factors of the disease. Among the main risk factors, low-density lipoproteins (LDLs), particularly in their oxidized status (ox-LDLs), are a causal determinant of atherosclerosis and CAD [17]. Ox-LDLs induce the expression of adhesion molecules in endothelial cells (ECs), mediate the inflammatory response, cause dysfunction and apoptosis of ECs, regulate the generation of reactive oxygen species, and are involved in vascular smooth muscle cell (VSMC) proliferation and apoptosis [18]. In this study we had 3 aims: 1) to identify miRNAs differentially expressed in human ECs and VSMCs, after exposure to ox-LDLs; 2) to analyze whether the miRNAs associated to ox-LDLs exposure, and others known to be associated to CAD, were differentially expressed in MI cases compared to healthy controls; and 3) to determine whether any of the miRNAs identified in objectives 1 and 2 could aid in predicting 10-year incident CAD in general population.

## Methods

### LDL cell treatment study

#### Human artery collection

ECs and VSMCs derived from healthy human renal arteries, from 3 individuals without CVD, were obtained from donors of the COLMAH collection (https://www.redheracles.net/plataformas/en_coleccion-muestras-arteriales-humanas.html). Vessels were divided into two pieces within the first 24h after extraction. One piece was placed in RNAlater (Ambion) for RNA extraction, and the other in Dulbecco’s modified Eagle’s medium (DMEM) for cell isolation.

#### EC and VSMC isolation

For EC culture, arteries were longitudinally opened, washed with PBS and incubated for 20-25 min with 1% collagenase type I in their endothelial side. Afterwards, ECs present in the collagenase solution were collected and washed twice with M199 medium containing 20% of fetal bovine serum (FBS). ECs were subsequently grown in 2 µg/ml fibronectin (Sigma) coated dishes, with human EC-specific medium EBM supplemented with EC growth factors (EGM-2, Lonza), and kept at 37 °C and 5% CO_2_. ECs were identified by their cobblestone morphology.

For VSMC culture, cells were isolated from the medial layer of the vessel after removal of ECs and the adventitia layer. The medial layer was cut in 1-3 mm pieces that were seeded in 35 mm Petri dishes treated with 2 % gelatin (Type B from bovine skin, Sigma) in DMEM with 20% FBS, penicillin-streptomycin (100 U/ml each), 5 μg/ml fungizone, and 2 mM L-glutamine (Lonza), and kept at 37 °C and 5% CO_2_. Migration and proliferation of VSMCs from the explants was evident within 7-15 days. Confluent VSMCs were trypsinized, seeded at 1/3 density, and cultured in control medium (MEM with 5% FBS, penicillin-streptomycin, fungizone, L-glutamine 5 μg/ml insulin, 2 ng/ml bFGF, and 5 ng/ml epidermal growth factor [19]).

#### LDL oxidation and cell treatment

Commercial human LDL (Prospec Pro-562) was diluted to 500 µg/ml in PBS (nLDL) and incubated with 5 µM CuSO_4_ for 30min, for moderately oxidized LDL (moxLDL). For highly oxidized LDL (hoxLDL), nLDL was incubated with 10 µM CuSO4 for 24h. The reaction was ended adding 200 µM EDTA to prevent further oxidation and then dialyzed (Slide-A-Lyzer Dialysis Cassete 3.5 MWCO, ThermoFisher 66110). The degree of LDL oxidation was assessed by measuring conjugated diene formation at 234 nm [20].

ECs and VSMCs at passages 4-8 were incubated with 50 µg/ml of nLDL, moxLDL, or hoxLDL for 24h. Then, cells were collected with Qiazol Lysis Reagent for RNA extraction.

#### Case-control study

An age- and sex-matched case-control study of 500 AMI cases and 500 controls was designed within the REGICOR (REgistre GIroni del COR) Study. Controls were randomly selected from the participants of a population-based cohort free of CAD symptoms during a 10-year follow-up. The cohort was recruited in the Girona province, in 2003-2005 [21]. AMI cases were randomly selected from the patients of the REGICOR AMI Registry, admitted consecutively with q-wave AMI in the reference hospital (Josep Trueta, Girona). The REGICOR AMI Registry was active during 1990-2009 in the same region as the REGICOR population cohort [22-24].

#### CV risk factor data and blood sample collection

The methods to obtain baseline data on CV risk factors for the 2005 REGICOR population cohort and for the REGICOR AMI Registry, have been detailed elsewhere [21-23]. Briefly, examinations in the 2005 REGICOR population cohort were performed by trained personnel using standard questionnaires and measurement methods. Collected/measured variables included age, sex, height, weight, blood pressure, fasting levels lipid levels (total cholesterol, triglycerides, and high- and low-density lipoprotein cholesterol [HDL-c, and LDL-c, respectively]), fasting glycaemia, and diagnosis and treatment of hypertension, hypercholesterolemia, and diabetes. Blood samples were collected after a 10-14h fasting and stored at -80°C. In the REGICOR AMI Registry, the same variables were obtained by trained personnel from clinical records. Blood samples were collected upon arrival to hospital and stored at -80°C.

#### Case-cohort study

A case-cohort study was designed also within the REGICOR population-cohort recruited in 2003-2005, with all 117 10-year incident CAD events, and a random subsample of the cohort paired with cases by age and sex (n=483).

#### Case-finding procedures and case-classification in the REGICOR population-cohort

Participants from the 2005 REGICOR population-cohort were re-examined or interviewed by telephone when they could not attend the appointment. AMI case investigation in the REGICOR Registry has been described previously [24]. Events of interest were identified with medical records and with the Mortality Register of Catalonia to identify fatal CAD events. Participants were classified as CAD incident cases if they had a discharge record suggestive of fatal or nonfatal AMI or angina (ICD-9 codes: 410, 411.0, 411.1, 412, 414, 429; and ICD-10 codes: I21-I25, including subtypes). Death certificates with codes 410-414 (ICD-9) or I20-I22, I24, and I25 (ICD-10) were selected for review of medical records and autopsy results. Events were classified by an expert committee following standardized guideline criteria [25]. Angina was defined according to the presence of symptoms and objective demonstration of ischemia or presence of coronary stenosis.

#### RNA extraction and quality control

Total RNA from LDL treated cells was isolated with miRNasey Mini kit (Qiagen) following manufacturer’s instructions. RNA extraction from serum samples, of the case-control and case-cohort studies, was performed with the MagMAX mirVana Total RNA Isolation kit (Thermo Fisher Scientific). RNA spike-in controls ath-miR159a and cell-miR-2 were added during the RNA extraction process from serum samples.

### miRNA expression

#### LDL cell exposure study

miRNA expression in LDL treated cells was examined using the miRNA 4.0 array (Thermo Fisher Scientific). RNA samples underwent polyadenylation and biotin-labelling with the Flash Tag Biotin HSR RNA Labeling Kit (P/N 703095, Rev.2) (Thermo Fisher Scientific). Biotin-labeled RNA samples were subsequently hybridized for 16h to the GeneChip miRNA 4.0 arrays, in a GeneChip Hybridization Oven 640. Arrays were washed and stained in a GeneChip Fluidics Station 450 and scanned in a GeneChip Scanner 3000 7G, using the Expression Wash, Stain and Scan User Manual (P/N 702731, Rev.3).

#### Case-control and case-cohort studies

RNA extracted from serum samples and from LDL treated cells (also examined with the miRNA array), underwent polyadenylation, adaptor ligation, and reverse transcription with the Taqman advanced miRNA cDNA synthesis kit (Thermo Fisher Scientific). cDNA samples were combined with the miR-Amp PCR master mix and transferred to a custom made OpenArray plates with the AccuFill instrument (Thermo Fisher Scientific). Custom OpenArray plates included 52 miRNAs: 24 identified in the LDL cell treatment study, 22 known to be associated with CAD, and 6 as housekeeping miRNAs (Supplementary material online, Table S1). OpenArray plates were run on the QuantStudio 12K Flex instrument (Thermo Fisher Scientific). qPCR amplification curves were examined with the Thermo Fisher Cloud software for Ct determination and for melting curve analysis. Observations with an AmpScore >1.1 and for which an exponential curve was present were included in the analysis.

### Statistical analysis

#### miRNA expression and selection criteria in the LDL cell treatment study

Background correction, normalization, and summarization of expression were performed with the RMA function of the aroma.affymetrix R package [27]. The RMA function uses a deconvolution method for background correction, quantile normalization, and the median polish algorithm for summarization of expression. To obtain differentially expressed miRNAs, linear models were fit by comparing RMA summarized expression between the following cell treatment conditions: moxLDL vs nLDL, and hoxLDL vs nLDL. Moderated t-statistics of differential expression were obtained by empirical Bayes moderation of the standard errors using the limma R package [28]. P-values were adjusted for multiple comparisons with the Benjamini-Hochberg correction [29].

miRNAs with a fold change ≥1.5 and a significant unadjusted p-value, in moxLDL vs nLDL or in hoxLDL vs nLDL, in at least 1 cell type (ECs or VSMCs), were selected for further analysis based also on the expression trend. The existence of a linear/quadratic trend in the miRNA expression between the conditions nLDL, moxLDL, and hoxLDL was assessed with linear models using generalized least squares. Trend p-values were adjusted with the Benjamini-Hochberg correction. miRNAs with a significant linear or quadratic trend in at least 1 cell type were selected.

#### Validation of the microarray miRNA expression

Similarity of miRNA expression obtained with the microarray and the OpenArray, was assessed with the Spearman correlation coefficient of RMA summarized expression and deltaCt normalized expression values, respectively.

#### miRNA expression analysis in the case-control and case-cohort studies

miRNAs with >90% of Ct missing values and individuals with >95% of Ct missing values were excluded from the analysis. miRNAs with <90% of missing Ct values were also excluded if they did not allow for plate effect correction. Cts were corrected for possible plate effects with the ComBat function of the sva R package [30]. Global normalization was performed by subtracting the Ct of each miRNA from the mean Ct of all miRNAs (Delta Ct).

In the case-control study the fold change was calculated to assess the association of each miRNA and case-control status. Fold change was defined as the 2~[-(DeltaCT cases – DeltaCT controls)]. Missing Ct values were censored at the maximum observed value for each miRNA. Means and p-values were computed with Kaplan-Meier and long rank test, respectively, to consider censored values.

In the case-cohort study, missing Ct values were removed from the analysis. An age- and sex-adjusted Cox regression model, taking into account the case-cohort design [31], was fitted to assess the association of each miRNA and time-to-CAD event. P-values were also adjusted for multiple comparisons with the Benjamini-Hochberg correction. The independent association of the significant miRNAs was examined with a Cox regression model further adjusted for classical risk factors (age, sex, smoking, cholesterol level, blood pressure, diabetes).

#### Predictive capacity of the identified miRNAs

In the case-cohort study, the contribution to the predictive capacity of the significant miRNAs, over classical risk factors, was analyzed with the C-statistic. The C-statistic was computed for a model with all classical risk factors, and including or not the most significant miRNAs identified in the case-control or in the case-cohort study. The increment of the C-statistic was calculated as previously described for case-cohort studies [32,33]. Reclassification was examined with categorical and continuous Net Reclassification Index (NRI), and with the integrated discrimination improvement (IDI) for the REGICOR CAD risk function [7]. For the categorical NRI, the cutoff point was 10%, which corresponds to the high risk category in the REGICOR CAD risk function. NRI and IDI confidence intervals were obtained by bootstrapping.

#### Pathway analysis

In the case-control study, gene targets of the differentially expressed miRNAs were obtained from the TargetScanHuman website release 7.2 [34]. Targets with a cumulative weighted context +++ score < -0.3 and with an aggregate score ≥ 0.3 were selected. Pathway analysis was performed using the hypergeometric test implemented in the ReactomePA R package [35]. P-values were adjusted for multiple comparisons with the Benjamini-Hochberg correction.

All statistical analyses were performed with the R software version 3.5.2. [36].

#### Ethics

The study was approved by the Parc de Salut Mar Ethics Committee (2015/6202/I). Participants signed a written informed consent.

#### Data availability statement

The data used in this study is available from the authors on reasonable request.

## Results

Characteristics of the study participants are presented in Supplementary material online, (Tables S2-S4).

### LDL cell treatment study

There were 21 miRNAs with differential expression that showed a linear or quadratic significant trend, in ECs or VSMCs, upon treatment with hoxLDL/moxLDL compared to nLDL (Figure 1, Supplementary material online, Tables S5 and S6). Of the 21 differentially expressed miRNAs, 4 were identified in both cell types (hsa-miR-3151-5p, hsa-miR-4669, hsa-miR-6831-5p, and hsa-miR-7107-5p). Hsa-miR-4669 and hsa-miR-6831-5p were down-regulated upon treatment with hoxLDL/moxLDL in both cell types. On the other hand, hsa-miR-3151-5p was down-regulated in ECs and upregulated in VSMCs, while hsa-miR-7107-5p did the opposite. The largest change in expression was observed for hsa-miR-197-3p in ECs (4 times more expressed in cells treated with nLDL) and for hsa-miR-122-5p in VSMCs (2.5 times more expressed in cells treated with hoxLDL).

**Figure 1.**
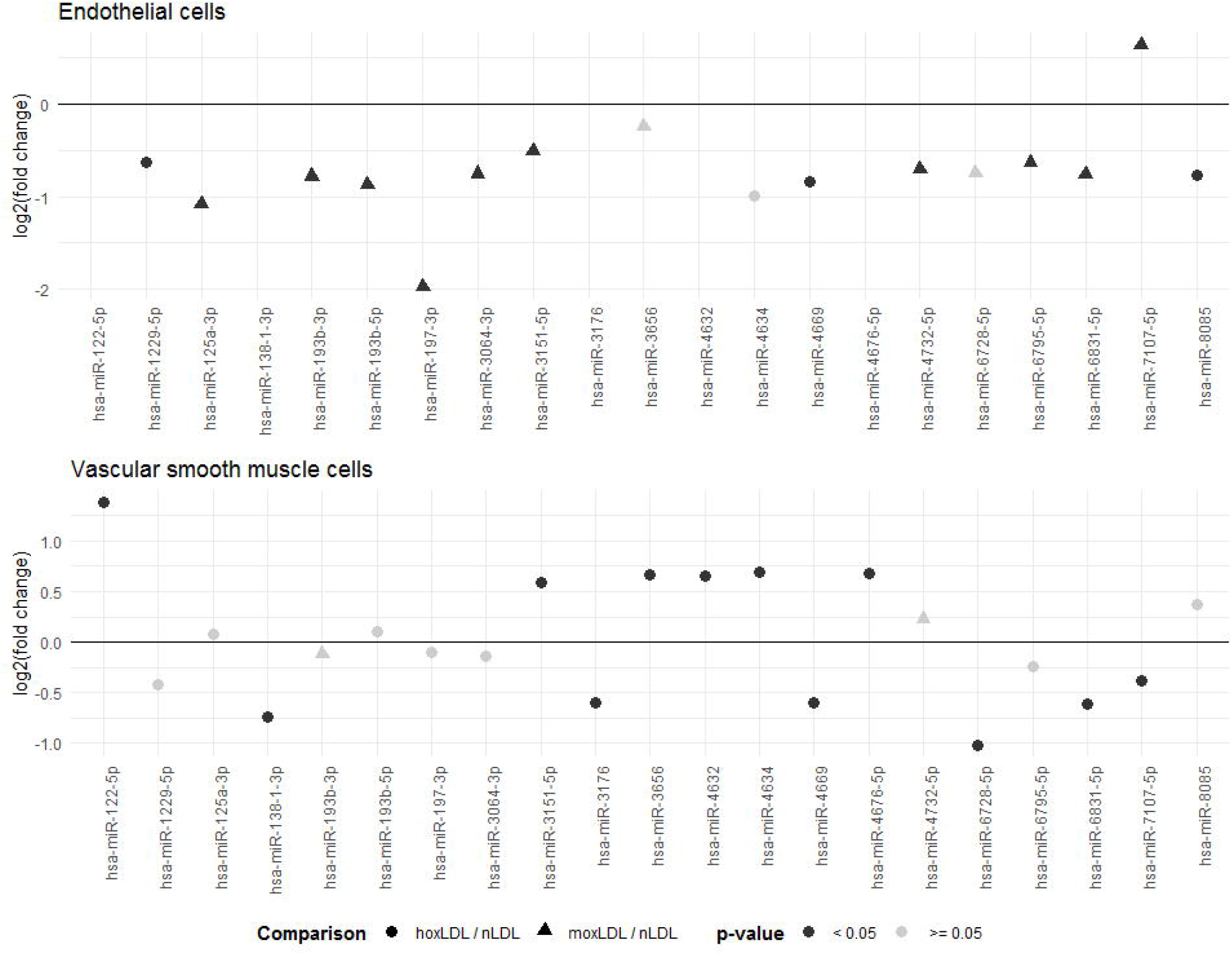
Differentially expressed miRNAs in endothelial and vascular smooth muscle cells which followed a linear or quadratic trend between treatment with native, moderately, and highly oxidized low density lipoproteins.

### Validation of miRNA expression in LDL treated cells

The higher the RMA summarized expression obtained with the microarray, the lower the DeltaCt obtained from the qPCR OpenArray (Supplementary material online, Figure S1). Correlation was significant for all samples (r = -0.475, p-value=1.22e^-06^) and by cell type (rEC =-0.36, rVSMC = -0.41, p-value<7e^-03^). miRNAs with a microarray RMA expression >3 (54%) were partially validated by qPCR (37%). Five out of 22 miRNAs differentially expressed in the microarray were detected by qPCR (hsa-miR-122-5p, hsa-miR-125a-3p, hsa-miR-193b-3p, hsa-miR-193b-5p, and hsa-miR-1229-5p).

### Case-control study

The analysis included 476 cases and 487 controls (Supplementary material online, Figure S2) with <95% of missing miRNA Cts, and 14 miRNAs with <75% of missing Ct values. Ofthe 14 candidate miRNAs, 12 were differentially expressed -and upregulated- in MI cases compared to controls (adjusted p-value<0.05) (Figure 2, Supplementary material online, Table S7). The largest difference in expression was observed for hsa-miR-499a-5p, hsa-miR-16-5p, and hsa-miR-133a-3p, which had an expression 118, 10, and 9 times higher, respectively, in MI cases compared to controls.

**Figure 2.**
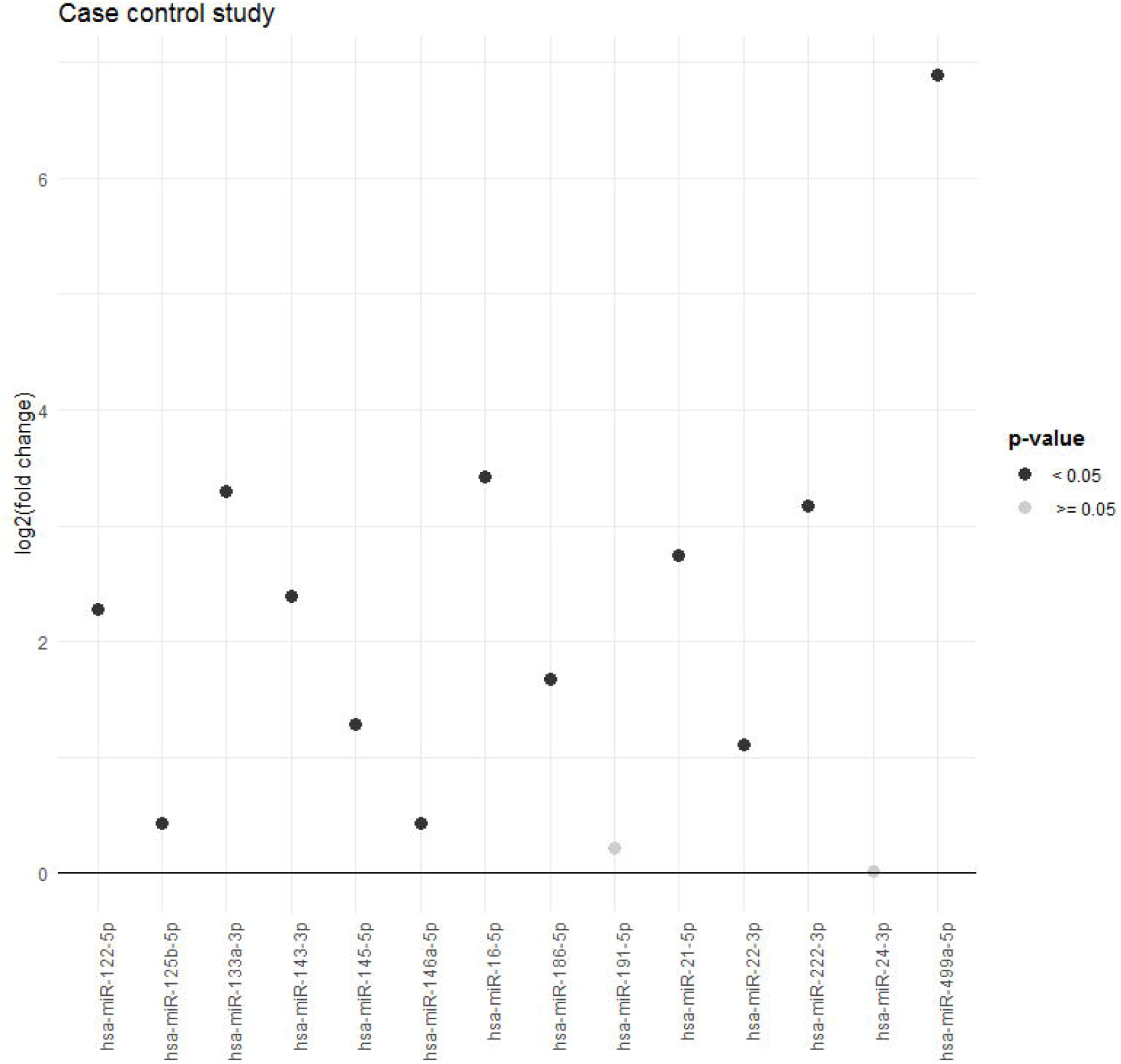
Differentially expressed miRNAs in acute myocardial infarction cases compared to controls.

TargetScan identified 7953 gene targets for the 12 upregulated miRNAs. Of those, the ones with higher binding efficacy and conservation (n=1431), were selected for pathway analysis. The identified targets were significantly associated with 53 pathways (Supplementary material online, Table S8). The 10 most significantly associated pathways included tyrosine, MAP kinase, toll like receptor, and cytokine signaling (Figure 3). In each pathway there were 19-72 gene targets.

**Figure 3.**
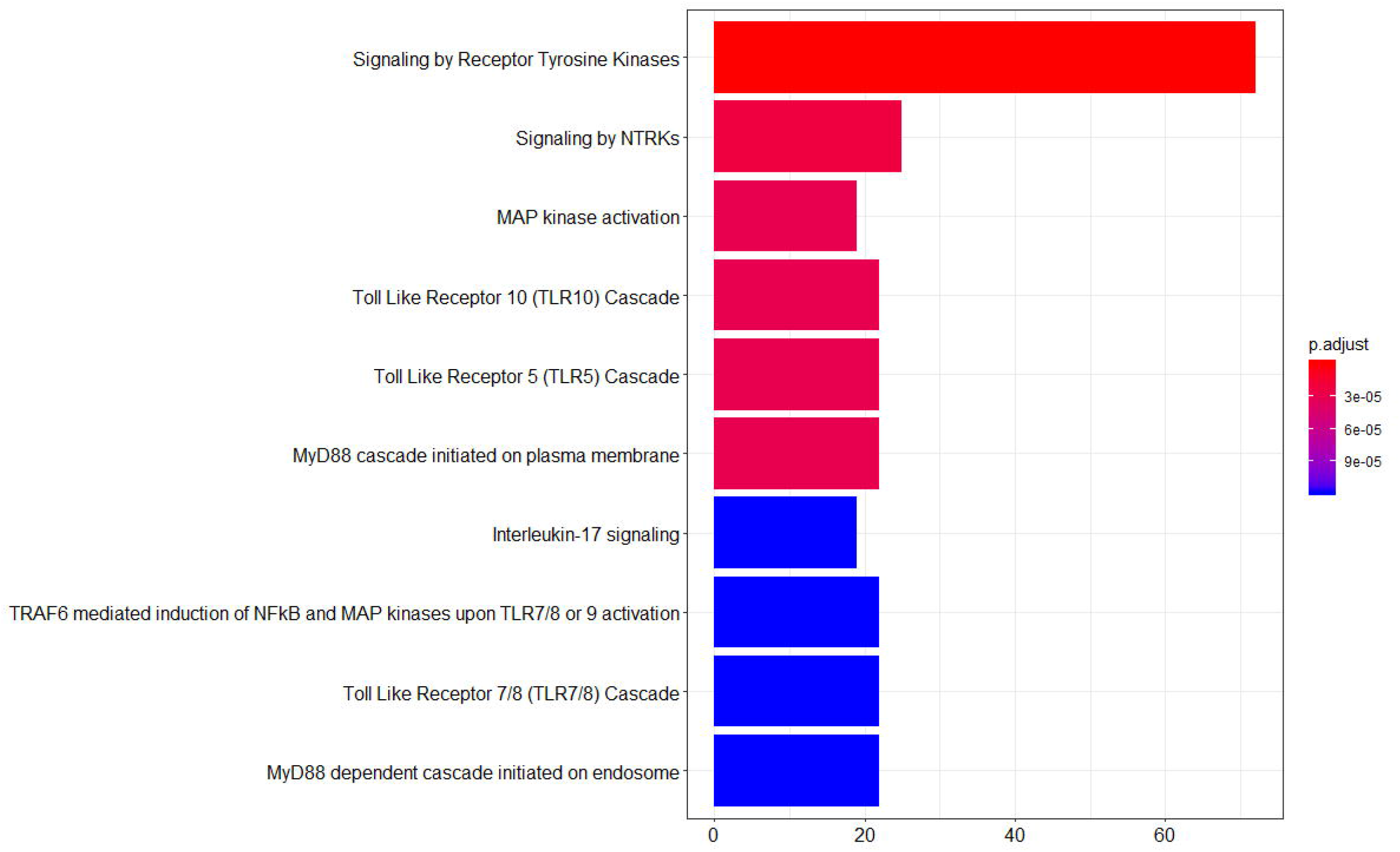
Top 10 most significant pathways associated with the most conserved and effective targets from the miRNAs differentially expressed in the case-control study.

### Case-cohort study

We analyzed 105 CAD cases and 455 individuals from the cohort (Supplementary material online, Figure S3), with ≤95% of missing miRNA Cts, and 14 miRNAs with ≤80% of missing Ct values. Only hsa-miR-143-3p was significantly associated with time-to-CAD incident events in the age- and sex-adjusted Cox model (hazard ratio=0.56 [95%Confidence Interval (CI) 0.38; 0.82], adjusted p-value=0.003) (Supplementary material online, Table S9). The association of hsa-miR-143-3p with CAD was independent of classical risk factors (p-value = 0.008).

The inclusion of the Delta Ct of hsa-miR-143-3p or of the 3 most significant miRNAs in the case-control study (hsa-miR-499-5p, hsa-miR-16-5p, hsa-miR-133a-3p), did not improve discrimination over classical risk factors (Figure 4, C-index). However, hsa-miR-143-3p, and its combination the 3 case-control miRNAs improved reclassification measures. Hsa-miR-143-3p improved continuous NRI (46.7% [95%CI 6.5; 88.7], p-value=0.026) and IDI (0.065 [95%CI 0.018; 0.113], p-value=0.007). And the combination of the 4 miRNAs improved IDI (0.303 [95%CI 0.116-0.490], p-value=0.001, respectively) (Figure 4, Continuous NRI and IDI).

**Figure 4.**
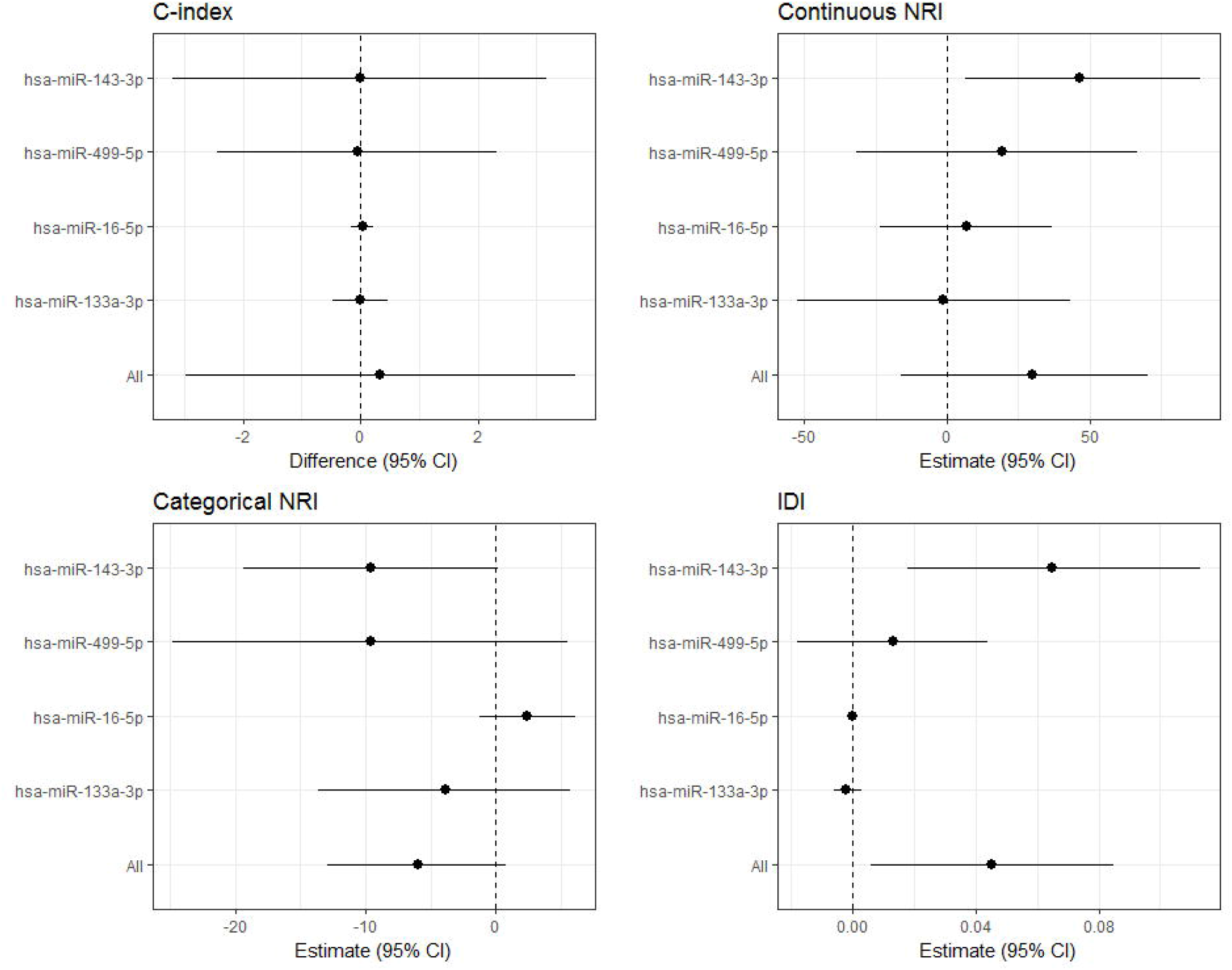
Predictive capacity of the miRNA differentially expressed in the case-cohort study, and of the 3 miRNAs most differentially expressed in the case-control study. Discrimination was measured with the C-index, and reclassification with the NRI (continuous and categorical) and with the IDI.

## Discussion

In this study, we identified 21 miRNAs differentially expressed upon treatment with ox-LDLs in EC and VSMCs. The miRNAs showing the largest expression change in VSMCs (hsa-miR-122-5p) was also upregulated in serum samples of MI cases compared to controls. Another 11 miRNAs, identified from the literature, were upregulated between MI cases and controls, particularly hsa-miR-499-5p. The predicted targets of the 12 upregulated miRNAs in the case-control study were associated with signaling pathways of tyrosine kinases, MAP kinases, and toll-like receptors. None of the miRNAs differentially expressed upon ox-LDL treatment in EC or VSMCs was associated with time-to-incident CAD event in the case-cohort study. However, hsa-miR-143-3p, a miRNA, known to be upregulated in AMI cases compared to controls, was independently and inversely associated with time-to-CAD incident event. The addition of hsa-miR-143-3p to classical risk factors improved significantly reclassification of population according to their CV risk, although discrimination remained unchanged.

After treating EC and VSMCs with ox-LDLs, the expression of 21 miRNAs significantly changed, when compared to nLDL treatment. A few of these miRNAs (hsa-miR-122-5p, hsa-miR-125a-3p, hsa-miR-193b-3p, hsa-miR-197-3p, hsa-miR-4632, and hsa-miR-7107-5p), were previously associated with ox-LDL related processes such as lipid metabolism, reactive oxygen species, and inflammation [37-42]. Of those, we were particularly interested in hsa-miR-122-5p, hsa-miR-125a-3p, and hsa-miR-193b-3p, as they were validated by qPCR. Hsa-miR-122-5p levels associated with lipoprotein components in general population, and with cholesterol and triglyceride levels in animal studies [37]. These associations were probably related to the effect of hsa-miR-122-5p on the regulation of genes involved in fatty acid synthesis and oxidation [43]. Hsa-miR-125a-3p was associated with pro-inflammatory cytokines and with cholesterol levels in obese individuals [38]. And hsa-miR-193b-3p was associated with production of reactive oxygen species in human fibroblasts [39]. Besides validating the association between ox-LDL related processes and hsa-miR-122-5p, hsa-miR-125a-3p, and hsa-miR-193b-3p, we also identified 2 novel miRNAs associated with ox-LDL treatment: has-miR-193b-5p and has-miR-1229-5p.

Hsa-miR-122-5p was not only upregulated in VSMCs upon treatment with ox-LDLs, but also in AMI cases compared to controls. This result adds up to the involvement of the LDL oxidation pathway in the response after an AMI [44]. In a recent meta-analysis, has-miR-122-5p was identified as upregulated in CAD cases compared to controls [45]. In addition, in hyperlipidemia patients, hsa-miR-122-5p was upregulated, and associated with presence and severity of CAD [46].

Besides hsa-miR-122-5p, we found 11 other upregulated miRNAs in AMI cases compared to controls (has-miR-16-5p, has-miR-21-5p, has-miR-22-3p, has-miR-125b-5p, has-miR-133a-3p, has-miR-143-3p, has-miR-145-5p, has-miR-146a-5p, has-miR-186-5p, has-miR-222-3p, and has-miR-499a-5p). Has-miR-499a-5p was the most upregulated miRNA, with a 118-fold expression change, an increase similar to that reported by 2 previous studies of AMI patients [47,48].

In accordance to our results, the 12 overexpressed miRNAs in the case-control study, were identified as significantly upregulated in individuals with AMI or CAD in previous studies [45, 49-52], or after cardiac ischaemia in animal models (miR-15 family) [53]. These miRNAs have been associated with 1) apoptosis in ECs and cardiomyocytes (has-miR-16-5p, has-miR-499a-5p, has-miR-21-5p, has-miR-22-3p, and has-miR-125b-5p) [54-57]; 2) proliferation and differentiation in VSMCs and cardiomyocytes (has-miR-145-5p, has-miR-143-3p, and has-miR-133a-3p) [49, 58, 59]; 3) angiogenesis (has-miR-222-3p, and has-miR-22-3p) [58]; 4) secretion of pro-inflammatory cytokines (has-miR-186-5p, has-miR-222-3p, and has-miR-22-3p) [58,59], 5) lipid accumulation (has-miR-186-5p) [59], and 6) fibrosis (has-miR-22-3p) [52]. By regulating these processes, the differentially expressed miRNAs are able to improve cardiac function (has-miR-22-3p), and to stimulate cardiac regeneration (has-miR-146a-5p), cardiac tissue contractibility and calcium handling (has-miR-21-5p) [54, 56].

To understand the processes regulated by the combination of the miRNAs upregulated in the case-control study, we performed pathway analysis with the most conserved and potentially effective targets. We found that the most associated pathways were related to signaling by MAP and other kinases, toll like receptors, and inflammatory cytokines. Kinase signaling pathways, including MAP and PI3K/Akt (both upregulated in our study), are associated with oxidation, angiogenesis, cell cycle, and inflammatory cytokines [61-63]. In addition, signaling trough toll like receptors triggers an inflammatory reaction [64]. The identified pathways suggest that the combined action of these 12 miRNAs regulates a subset of the individual miRNA functions, particularly inflammation. Taking into account that the most significant pathways identified in our study have already been associated with AMI [64-67], this group of miRNAs can explain, at least in part, the molecular response after AMI. And thus, these 12 miRNAs could be used to improve the diagnosis of AMI, and the prognosis prediction in AMI patients. On the one hand, has-miR-499-5p and has-miR-133a-3p showed high sensitivity and specificity values for the diagnosis of AMI in meta-analyses [68, 69]. Regarding prognosis, has-miR-499-5p was associated with reduced systolic function, and increased risk of death or heart failure after MI [70]. While has-miR-21-5p, has-miR-143-3p, and has-miR-15a-5p (a member of the miR-15 family that also includes miR-16), were upregulated in CAD patients with recurrent cardiovascular events [58].

Our case-cohort study identified has-miR-143-3p as the only miRNA associated with time-to-coronary events independently of classical risk factors. Moreover, adding has-miR-143-3p, alone or in combination with the 3 most differentially expressed miRNAs in the case-control study, to the commonly used risk factors improved reclassification measures. Only the Hunt study has analyzed miRNA expression in healthy individuals to predict coronary events [15]. The Hunt investigators identified 5 miRNAs that improved reclassification and discrimination. However, these 5 miRNAs were different from the ones identified in the present study, probably due to the different methodology applied. While the Hunt investigators used a small panel of serum miRNAs (n=179), we focused on the miRNAs associated with ox-LDL treatment from a larger miRNA panel (n=2,578), and on previously reported miRNAs. Although the results differ, both studies suggest that new biomarkers such as miRNAs could be useful to improve CAD risk prediction in general population. While has-miR-143-3p was upregulated in AMI cases compared to controls, a higher expression in healthy individuals was in contrast associated with a reduced risk of having an incident CAD event. This result reflects the complex time- and cell type-specificity of miRNA regulation. miR-143-3p is involved in the regulation of the phenotype and function of EC and VSMCs [71, 72], an in the initiation of the inflammatory response in the myocardium [73]. The increased expression of has-miR-143-3p in AMI patients is probably a response to the acute event, as observed in ischemic stroke patients, in which its levels decreased after the acute phase [74]. This acute upregulation of has-miR-143-3p could mediate the initiation of the inflammatory response that is triggered after AMI [75]. In addition, an increase in has-miR-143-3p expression could also stimulate vessel stabilization through the transfer of this miRNA from VSMCs to ECs [76]. On the other hand, miR-143 promotes differentiation and represses proliferation of VSMCs [71], and was downregulated in animal models of atherosclerosis [71], and in CAD patients [77].

These results agree with our observation of a protective effect of has-miR-143-3p on the CAD risk in healthy individuals. miR-143-3p, which is highly expressed in the normal vascular wall, would maintain the differentiated state of VSMCs, while its downregulation would be associated to a proliferating and less differentiated phenotype. Our study is the first analyzing the contribution of miRNAs induced by ox-LDLs to the identification of AMI cases and to CAD risk prediction in general population. We validated miRNAs associated to ox-LDLs related processes, discovered new miRNAs expressed after ox-LDLs treatment in EC and SMCs, validated miRNAs associated with AMI, and identified miRNAs associated with CAD incidence in general population. Our study has also some limitations that should be considered. In addition to validating previously identified miRNAs in CAD, we wanted to analyze the contribution of ox-LDL induced miRNAs, to the miRNA response in AMI, and to the miRNA expression in general population to predict future CAD. However, among the miRNAs associated with ox-LDL treatment, only 1 was dysregulated in AMI cases compared to controls, and none was related to CAD incidence. This result is probably related to the different miRNA response to ox-LDLs that occurs *in vivo* and *in vitro*. We palliated this limitation by including all previously miRNAs known to be associated to CAD. Another limitation is that we could only validate 5 out of 21 ox-LDL induced miRNAs in the study with EC and SMCs. The low validation could be associated with a lower miRNA concentration in serum compared to cell extracts.

In conclusion, we identified 2 novel miRNAs associated with ox-LDLs (has-miR-193b-5p and has-miR-1229-5p). We also found 4 miRNAs that improve the reclassification capacity of the REGICOR CAD risk function of general population based on their predicted CAD risk (has-miR-143-3p, and its combination with has-miR-16-5p, hsa-miR-133a-3p, and hsa-miR-499-5p). Our findings provide more knowledge on the response to ox-LDLs at the cellular level, and also candidate biomarkers to improve CAD risk prediction in general population.

## Data Availability

The data used in this study is available from the authors on reasonable request

